# Light Convolutional Neural Network to Detect Chronic Obstructive Pulmonary Disease (COPDxNet): A Multicenter Model Development and External Validation Study

**DOI:** 10.1101/2025.07.30.25332459

**Authors:** AKM Shahariar Azad Rabby, Muhammad F. A. Chaudhary, Pratim Saha, Venkata Sthanam, Arie Nakhmani, Chengcui Zhang, R. Graham Barr, Jessica Bon, Christopher B. Cooper, Jeffrey L. Curtis, Eric A. Hoffman, Robert Paine, Abhilash Kizhakke Puliyakote, Joyce D. Schroeder, Jessica C. Sieren, Benjamin M. Smith, Prescott G. Woodruff, Joseph M. Reinhardt, Surya P. Bhatt, Sandeep Bodduluri

## Abstract

**Background:** Approximately 70% of adults with chronic obstructive pulmonary disease (COPD) remain undiagnosed. Opportunistic screening using chest computed tomography (CT) scans, commonly acquired in clinical practice, may be used to improve COPD detection through simple, clinically applicable deep-learning models. We developed a lightweight, convolutional neural network (COPDxNet) that utilizes minimally processed chest CT scans to detect COPD.

**Methods:** We analyzed 13,043 inspiratory chest CT scans from the COPDGene participants, (9,675 standard-dose and 3,368 low-dose scans), which we randomly split into training (70%) and test (30%) sets at the participant level to no individual contributed to both sets. COPD was defined by postbronchodilator FEV /FVC < 0.70. We constructed a simple, four-block convolutional model that was trained on pooled data and validated on the held-out standard- and low-dose test sets. External validation was performed using standard-dose CT scans from 2,890 SPIROMICS participants and low-dose CT scans from 7,893 participants in the National Lung Screening Trial (NLST). We evaluated performance using the area under the receiver operating characteristic curve (AUC), sensitivity, specificity, Brier scores, and calibration curves.

**Findings:** On COPDGene standard-dose CT scans, COPDxNet achieved an AUC of 0.92 (95% CI: 0.91 to 0.93), sensitivity of 80.2%, and specificity of 89.4%. On low-dose scans, AUC was 0.88 (95% CI: 0.86 to 0.90). When the COPDxNet model was applied to external validation datasets, it showed an AUC of 0.92 (95% CI: 0.91 to 0.93) in SPIROMICS and 0.82 (95% CI: 0.81 to 0.83) on NLST. The model was well-calibrated, with Brier scores of 0.11 for standard-dose and 0.13 for low-dose CT scans in COPDGene, 0.12 in SPIROMICS, and 0.17 in NLST.

**Interpretation:** COPDxNet demonstrates high discriminative accuracy and generalizability for detecting COPD on standard- and low-dose chest CT scans, supporting its potential for clinical and screening applications across diverse populations.

## INTRODUCTION

Chronic obstructive pulmonary disease (COPD) is the third leading cause of death worldwide and was responsible for over 3 million deaths in 2019.^1–4^ COPD is estimated to affect nearly 392 million individuals globally, and 27 million in the United States of America (USA).^2,5,6^ Of these, approximately 95% worldwide, and 70% in the USA remain undiagnosed.^7–9^ Spirometry, currently required for diagnosis, is underutilized in many primary care settings due to limited availability and a lack of trained personnel, leading to underdiagnosis and delayed treatment.^10–12^ An attractive alternative approach to detect COPD, allowing for timely intervention and potentially improving patient outcomes, is the use of chest CT scans. Over 14 million chest CT scans are acquired annually in the USA alone.^13–15^ However, although machine- and deep-learning methods have shown promise in detecting COPD, existing approaches are not fully automated and often require manual intervention. For instance, Gonzalez *et al*. proposed a two-dimensional (2D) convolutional neural network (CNN) to detect COPD that was trained on a montage of four chest CT slices selected manually.^16^ Similarly, Tang *et al*.’s deep residual CNN required manual selection of 3D regions of interest for training.^17,18^ Recently, we used chest CT radiomics to detect COPD in both standard and low-dose CT scans^19^, an advance given the availability of the latter due to lung cancer screening. However, our method also requires feature extraction and selection for optimal performance. All these methods have demonstrated high accuracy in detecting COPD, but their reliance on expert oversight for handcrafted features and specific region selection limits their clinical applicability.

Here, we aimed to overcome these limitations by developing a lightweight CNN capable of analyzing minimally processed standard- and low-dose inspiratory chest CT scans to detect COPD. To develop and evaluate our model, we analyzed standard- and low-dose chest CT scans from large multicenter cohort studies.^20^

## METHODS

### Study Design

We developed our lightweight CNN, termed COPDxNet, in an initial training and validation approach on inspiratory standard-dose and low-dose chest CT scans, followed by external validation on two additional cohorts. The pooled training set comprised both standard and low-dose CT scans; we then evaluated the model separately on each dosage type to assess performance across different radiation doses. The ground truth for the diagnosis of COPD was defined spirometrically, as the ratio of forced expiratory volume in one second (FEV_1_) to the forced vital capacity (FVC) of < 0.70. We used the updated Transparent Reporting of a multivariable prediction model for Individual Prognosis Or Diagnosis (TRIPOD + AI) guidelines for reporting the development and validation of our COPD detection method.^21,22^

### Study Participants & CT Imaging

The training and test sets used data from participants in phases one (standard dose CTs, *n* = 9,675; at enrollment) and three (low-dose CTs, *n* = 3,368; follow-up at 10 years) of the Genetic Epidemiology of COPD (COPDGene) study.^20^ COPDGene is a large multicenter prospective cohort study being conducted at 21 clinical centers across the USA.^20^ Briefly, COPDGene enrolled adults aged between 45 and 80 years, who were current or former smokers with a smoking history of at least 10 pack-years.^20^ Participants underwent pre- and post-bronchodilator (BD) spirometry at each study visit, and we used post-BD values to define COPD. By design, COPDGene recruited approximately one-third of self-reported Black individuals.

COPDGene participants also underwent inspiratory chest CT scans that were acquired at total lung capacity.^20,23^ Standard-dose CT scans were acquired at enrollment on visit 1, and low-dose scans were acquired at visit 3 of the study, approximately 10 years later. For training and validation of COPDxNet, the combined dataset was randomly split into non-overlapping train (70% of scans, 6,775 standard-dose and 2,391 low-dose) and test (30% of scans, 2,900 standard-dose and 977 low-dose) sets (see **Fig. 1B)**. We stratified the pooled dataset by participant to ensure that scans from the same individual did not appear in both training and testing sets, thereby preventing data leakage and potential model bias. For more details on the COPDGene inclusion and exclusion criteria, please refer to **Table S1** in the supplementary appendix.

**Figure 1.**
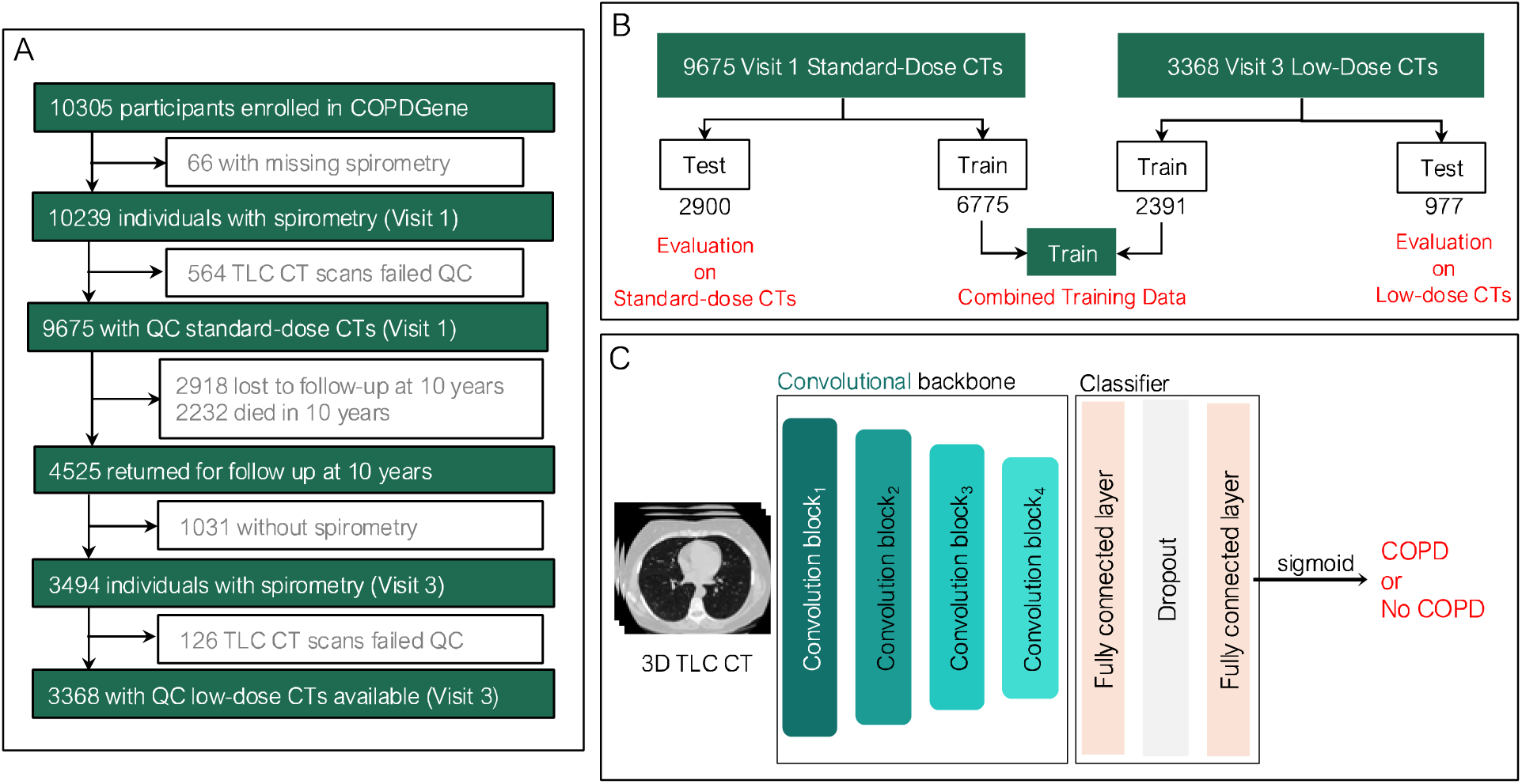
**Overview of the study design, model architecture, and external validation**. Flow diagram indicating the number of COPDGene participants available for analysis from visits 1 and 3 (Panel A). Model training and evaluation datasets with standard and low-dose chest CT scans. The model was trained on a single dataset consisting of both standard and low-dose scans. After training, the model was evaluated separately on standard and low-dose scans (Panel B). The four-block architecture of COPDxNet was used to process 3D inspiratory chest CT scans to detect COPD (Panel C). CT = Computed tomography, TLC = Total lung capacity, COPD = Chronic Obstructive Pulmonary Disease, QC = quality control.

For external validation, we analyzed data from two other large multicenter cohort studies. The Subpopulations and Intermediate Outcome Measures in COPD Study (SPIROMICS), ongoing at 14 clinical centers in the USA, initially enrolled 2,981 participants aged 40 – 80 years, including individuals with varying severity of COPD, smokers without airflow obstruction, and non-smoking controls.^24^ SPIROMICS Participants underwent pre- and post-BD spirometry and high-resolution chest CT scans at full inspiration.^25^

We additionally used data from the National Lung Screening Trial (NLST), which enrolled over 53,000 heavy smokers aged 55 – 74 years and randomized them to either low-dose chest CT or chest X-ray screening.^26^ For this study, we analyzed a subset of 7,893 participants from the CT arm who underwent pre-BD spirometry and low-dose CT at enrollment with slice thickness no greater than 2.0 mm, made available through the University of Alabama at Birmingham (UAB) Hospitals.^26^

All COPDGene, SPIROMICS, and NLST study procedures were conducted following the guidelines of the Declaration of Helsinki. Written informed consent was provided by all participants, and the protocols were approved by the Institutional Review Boards (IRBs) of each participating study center.

### Image Preprocessing

Image preprocessing steps included isotropic resampling of the images to have a voxel of size 0.5 mm^3^, clipping image intensities between –1024 and 400 Hounsfield Units (HU), and normalization of voxel intensities between –1 and 1. The processed images were then downsampled to have a fixed size of 128 × 128 × 64 voxels before training to include the entire lung.

### Light Convolutional Model Architecture – COPDxNet

We developed COPDxNet, a simple, four-block, 3D CNN architecture (see **Fig. 1C**). Each convolutional block was composed of a 3D convolutional layer with the rectified linear unit (ReLU) activation function, followed by max pooling and batch normalization. Dropout layers were inserted between convolutional blocks to help prevent overfitting. (see **Fig. 1C**). The model input was a batch of 3D images resized to 128 × 128 × 64. The first block was designed to learn 64 feature maps, which were successively increased to 256 feature maps in the final convolutional block. At the end of the convolutional backbone, a classification head composed of two fully connected layers was used for binary classification (see **Fig. 1C**). The feature maps obtained from the last convolutional block were passed to a fully connected layer with 512 perceptron units followed by a ReLU activation function. The final layer produced a scalar output, which was passed through a sigmoid activation function to predict the likelihood of COPD. The dropout rate at each stage was set to 0.5. The network was implemented using the open-source framework TensorFlow 2.0.

### Training Details

We trained our model using the binary cross-entropy loss function that was minimized by the Adam optimizer. To avoid premature convergence into a local minimum, we used an exponential learning rate decay, which automatically reduced the learning rate during training if the validation loss did not improve after a certain number of epochs. The decaying schedule was defined by an initial learning rate of 10^-4^, decay steps of 10^5^, and a decay rate of 0.96. More details on model training are provided in the Supplementary Appendix.

### Statistical Analysis

We evaluated the discriminative accuracy of COPDxNet using the area under the receiver operating characteristic curve (AUC), sensitivity, and specificity, computing the latter two by selecting a threshold that maximized the Youden’s index.^27^ To assess likelihood calibration, we employed calibration plots between the observed and the predicted probabilities and used Brier scores to evaluate model calibration quantitatively. To improve model calibration, we applied temperature scaling, a single-parameter variation of Platt scaling, across three timesteps.^28^ We also conducted a stratified analysis to evaluate the performance of COPDxNet across sex, smoking status, and different CT scanner types.

## RESULTS

### Characteristics of the Datasets for Training and Validation

Participant characteristics from the first and third visits of COPDGene, stratified by training and testing sets, are shown in **Table 1**. As anticipated, the mean age for the low-dose train and test sets was approximately 10 years higher than the mean age at enrollment. Both train and test sets were fairly balanced for sex (see **Table 1)**. In pooled training set 3,003 (44%) individuals with standard-dose CTs at visit 1 were labeled as COPD (defined by the FEV_1_/FVC < 0.70), while 990 (41%) individuals with low-dose CTs at visit 3 had COPD (see **Table 1**). We evaluated our model on 2,900 individuals with standard-dose and 977 individuals with low-dose CT scans separately, of whom, 1,264 (44%) and 383 (39%) had spirometric airflow obstruction, respectively (see **Table 1**). The training and testing sets from COPDGene were partitioned at the participant level to prevent data leakage from overlapping subjects.

**Table 1:**
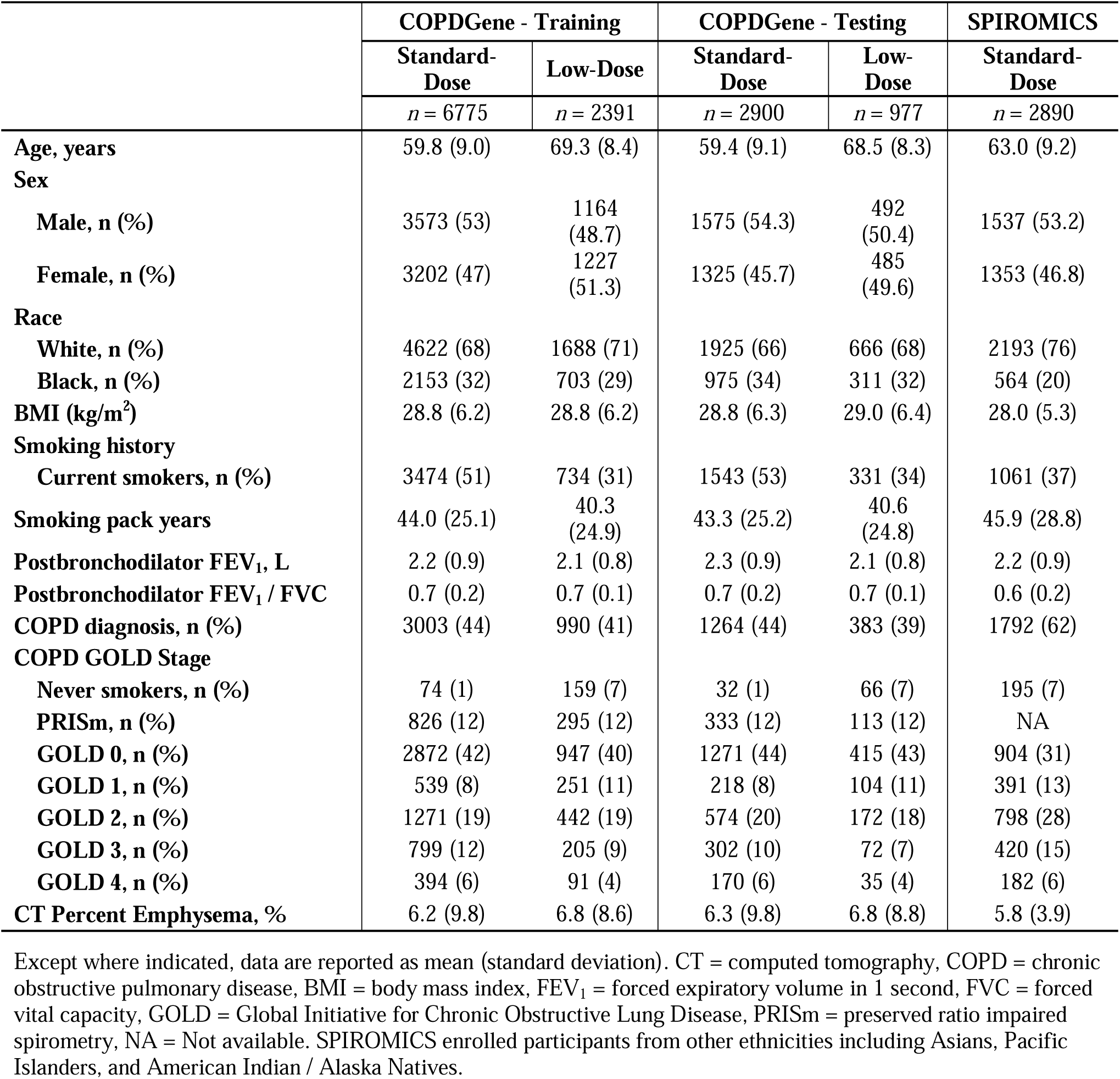
Participant characteristics from training and validation cohorts from COPDGene and SPIROMICS stratified by the acquisition of standard and low-dose chest CT scans.

Of 2,981 participants in SPIROMICS, eight withdrew consent, and 83 had missing imaging or spirometry data. For the remaining 2,890 participants, the mean age was 63.0 (9.2) years, with 1,537 males (53%). A total of 1,792 (62%) individuals had airflow obstruction, and 1,061 (37%) individuals were current smokers (see **Table 1**). NLST participants had a mean age of 61.7 (5.1) years, with 4,376 (55%) males, and 3,957 (50%) current smokers (see **Supplemental Table S2** of online appendix).

### Performance on Standard- and Low-Dose CT Scans in COPDGene

In the held-out test set, the discriminative accuracy of COPDxNet for detecting COPD on standard-dose chest CT scans was 0.92 (95% CI: 0.91 to 0.93), with sensitivity of 80.22% (95% CI: 75.83% to 82.65%), and specificity of 89.36% (95% CI: 87.87% to 93.06%) (see **Fig. 2A**).

**Figure 2.**
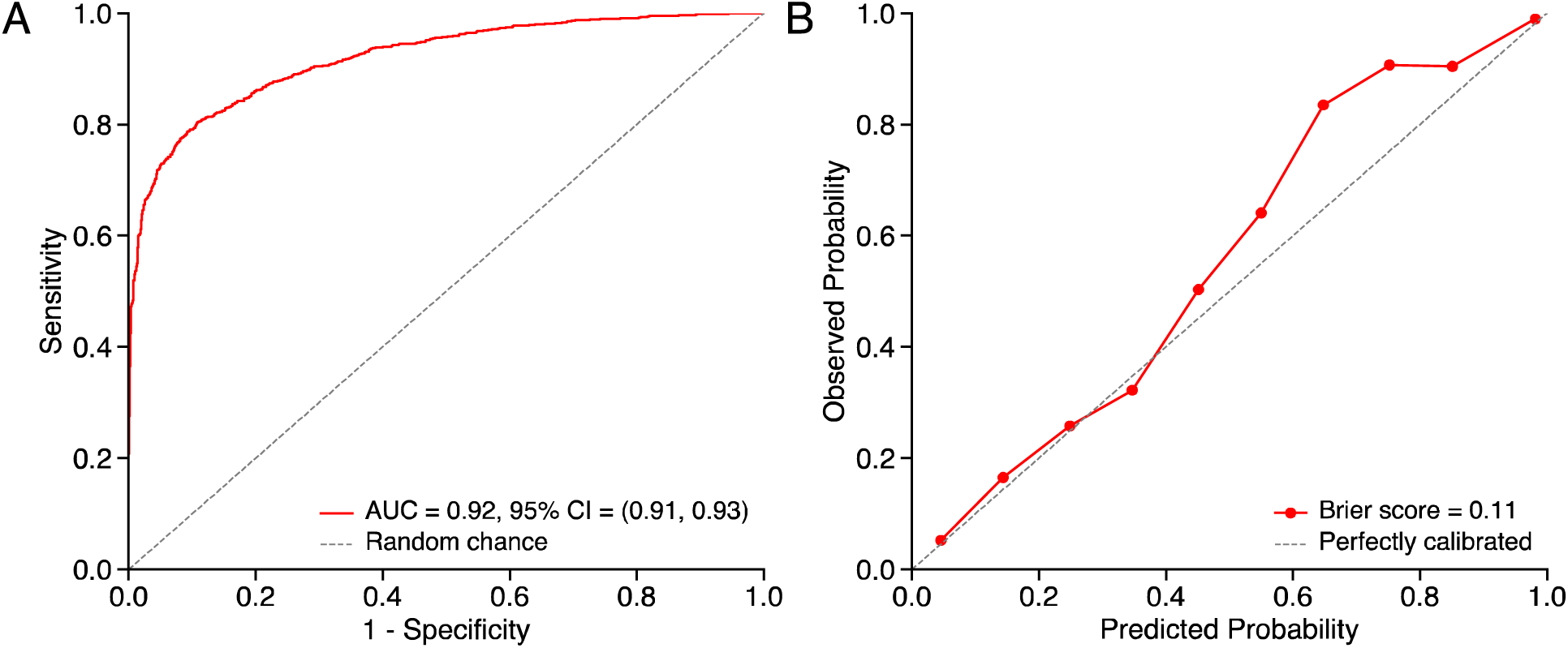
Performance evaluation on standard-dose chest CT scans (*n* = 2,900) using receiver operating characteristic curve (ROC) (Panel A) and calibration plots (Panel B).

The model displayed good performance on standard dose CTs (AUC 0.92, CI 0.92 to 0.93) and was well-calibrated, with a Brier score of 0.11 (95% CI: 0.10 to 0.12) (see **Fig. 2B**).

In the held-out low-dose test set, the discriminative accuracy of COPDxNet for the detection of COPD was high with an AUC of 0.88 (95% CI: 0.86 to 0.90) (see **Fig. 3A**), sensitivity of 81.46% (95% CI: 71.39% to 84.73%), and specificity of 81.48% (95% CI: 79.49% to 91.87%). The Brier score was 0.13 (95% CI: 0.12 to 0.15), which indicates reliable model calibration and reliable agreement between the predicted and observed probabilities (see **Fig. 3B)**.

**Figure 3.**
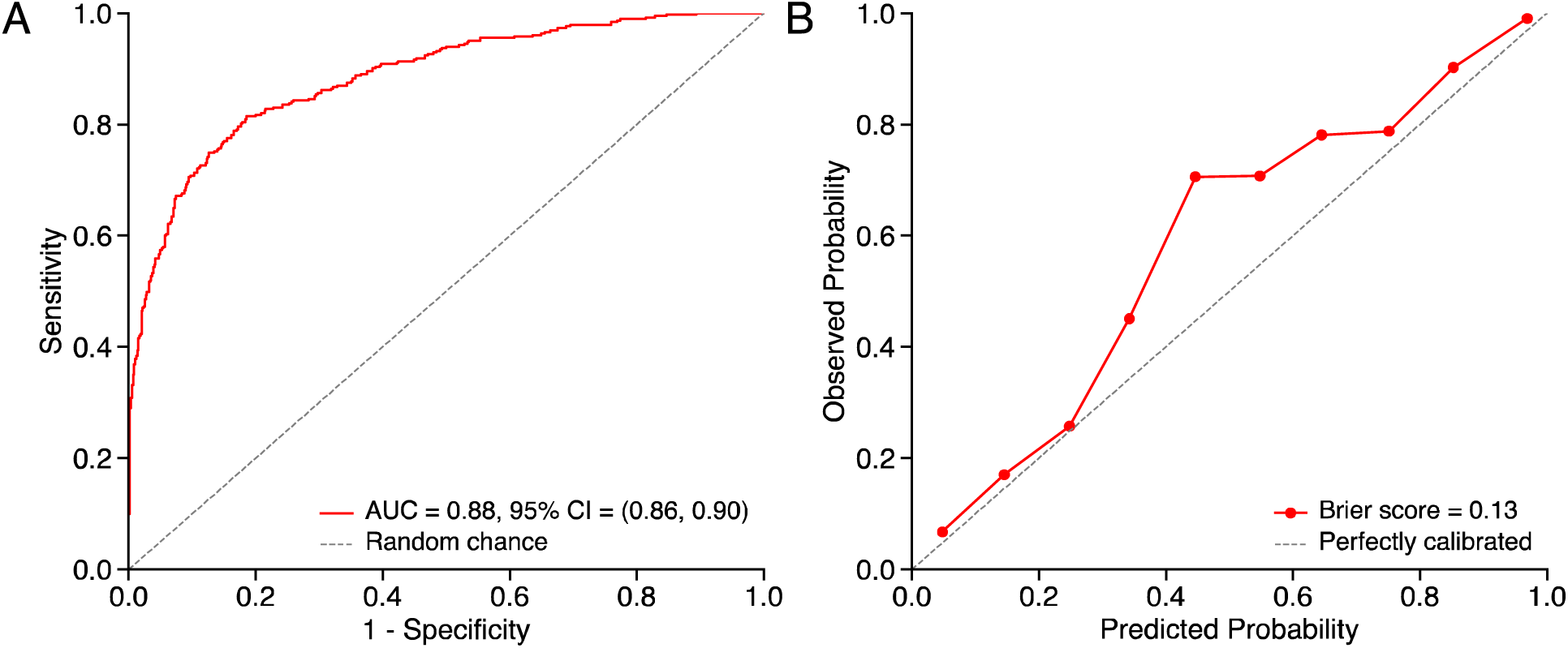
Evaluation of COPDxNet on low-dose chest CT scans (*n* = 977) using ROC curve (Panel A) and calibration plots (Panel B).

### Performance on SPIROMICS and NLST

When the COPDxNet model, trained on COPDGene, was applied to standard-dose chest CT scans from SPIROMICS, it generalized well with a similar AUC of 0.92 (95% CI: 0.91 to 0.93), sensitivity of 82.31% (95% CI: 80.90% to 83.80%), and specificity of 85.66% (95% CI: 83.80% to 87.40%) (see **Fig. 4A**). The predicted probabilities were well-calibrated in SPIROMICS as well with a Brier score of 0.12 (95% CI 0.11 to 0.13) (see **Fig. 4B**).

**Figure 4:**
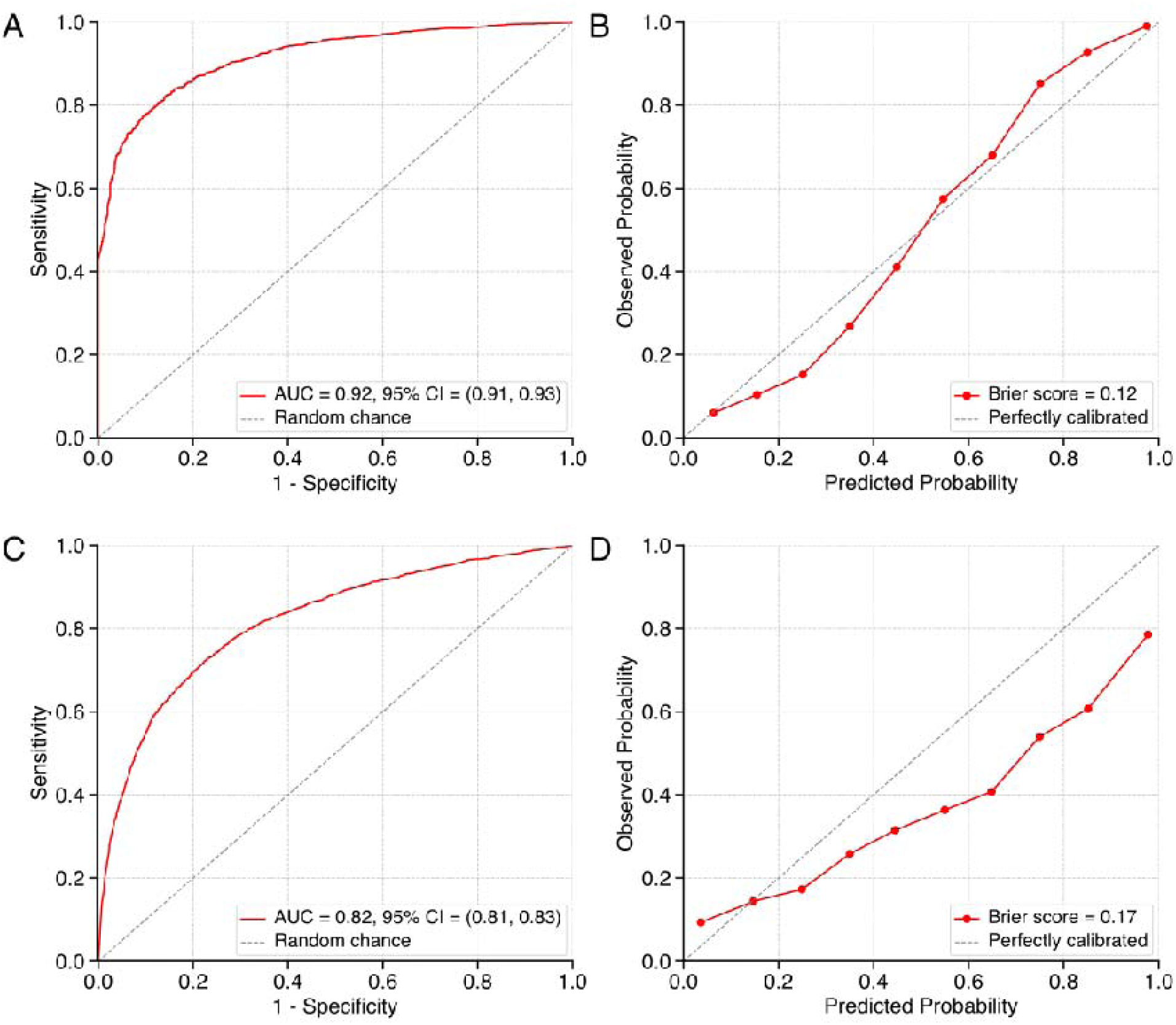
Discriminative performance evaluation of COPDxNet on external validation cohorts from SPIROMICS (Panel A) and NLST (Panel C). Calibration plots for detecting COPD in SPIROMICS (Panel B) and NLST (Panel D).

COPDxNet had a high discriminative accuracy on the low-dose, external validation test set from NLST: AUC of 0.82 (95% CI: 0.81 to 0.83), sensitivity of 70.60% (95% CI: 69.20% to 72.00%), and specificity of 78.97% (95% CI: 78.00% to 79.90%) (see **Fig. 4C**). The model was well calibrated with a Brier score for predicted probabilities of 0.17 (95% CI: 0.17 to 0.18) (see **Fig. 4D**).

### Sensitivity Analyses

In sensitivity analyses, we tested the performance of COPDxNet in several subgroups of COPDGene data. The discriminative accuracy of COPDxNet was high in men (AUC 0.92, 95% CI: 0.91 to 0.93 in standard-dose and 0.88, 95% CI: 0.85 to 0.90 in low-dose) and in women (0.92, 95% CI: 0.91 to 0.93 and 0.89, 95% CI: 0.86 to 0.91) (see **Supplemental Fig, S1**). The discriminative accuracy was high in both current smokers (standard-dose: 0.89, 95% CI: 0.88 to 0.91 and low-dose: 0.87, 95% CI: 0.84 to 0.91) and former smokers (standard-dose: 0.94, 95% CI: 0.93 to 0.95 and low-dose: 0.89, 95% CI: 0.86 to 0.91) (see **Supplemental Fig. S2)**. Our model performed well across different CT scanner types with AUCs between 0.88 and 0.94 (see **Supplemental Fig. S3**).

## DISCUSSION

We demonstrated that COPDxNet has a high discriminative accuracy for detecting COPD on both standard-dose and low-dose chest CT scans with minimal need for image preprocessing. The high volume of chest CT scans acquired in clinical practice, especially low-dose scans for lung cancer screening, offers an opportunity to screen for previously undetected COPD.^13,14,26^

COPDxNet has significant advantages over the several previously reported algorithms for COPD detection using machine learning and deep learning techniques.^16,17,19,29^ These algorithms were limited by the need for significant preprocessing or manual input for slice selection. A 2D CNN that used a montage of four selected slices from axial, coronal, and sagittal views was associated with an AUC of 0.86 for the detection of COPD. This model required significant manual oversight to select the slices, which may reduce its clinical applicability. Du et al. developed a CNN that used multiple 2D CT slices and reported an accuracy of 88.6%.^30^ The CT slices were obtained from different views (ventral and dorsal) of a 3D CT volume, and majority voting was used to label COPD. This approach also requires the preselection of 2D slices for detecting COPD. A 3D CNN architecture by Ho and colleagues used parametric response mapping measurements to detect COPD.^31^ This approach requires an additional expiratory chest CT scan, which is not routinely acquired in clinical practice and may increase exposure to radiation as well as costs. This approach also relies on accurate image registration of the inspiratory and expiratory chest CT scans. Furthermore, none of these models were evaluated on low-dose CT scans. COPDxNet is associated with higher discriminative accuracy than prior models and has the advantages of minimal preprocessing and the use of a lightweight 3D CNN architecture which efficiently processes the entire 3D CT volume using a simple four-block backbone. Importantly, unlike most studies that did not investigate model generalization to large external validation cohorts, COPDxNet detected airflow obstruction with a high discriminative accuracy on standard dose CT scans from SPIROMICS. It is also worth noting that although the SPIROMICS scans were acquired at a higher dose than typical low-dose chest CT protocols, the radiation exposure was modulated based on body mass index (BMI) and remained significantly lower than the standard-dose scans used in Phase 1 of the COPDGene study.

With a growing drive to lower radiation exposure, chest CT scans are being acquired at increasingly lower doses and are being protocolled more frequently.^26,32,33^ COPD remains severely underdiagnosed within the general population and can benefit from the widespread availability of these low-dose scans. However, it is important that COPD detection models perform well on low-dose chest CT scans. Low-dose scans differ from standard dose scans in that they are generally associated with higher estimates of emphysema and airway disease.^34,35^ Such differences are a challenge for the development and generalization of automated COPD detection models. Tang *et al*. used a 2D residual CNN architecture to develop a COPD detection model that achieved an AUC of 0.89 on low-dose CT scans but required a preselection of axial image regions.^17^ To localize the region of interest, their approach required the additional step of aligning the input CT scans to a reference non-COPD CT scan. This step can be a significant bottleneck to the detection pipeline. We recently developed a radiomics framework for detecting COPD on standard and low-dose scans.^19^ While the model achieved a high discriminative accuracy, it required feature extraction and selection to refine its performance. In addition to evaluating our model on low-dose chest CT scans from the development cohort (COPDGene), we externally evaluated its discriminative accuracy on low-dose chest CT scans from NLST.

Our study has several strengths. We developed our model using scans acquired at 21 different clinical centers across the USA. Model training on such a diverse dataset, acquired across varying COPD severities, with a high representation of women and individuals of Black race, increases its generalizability. We trained our model on a combined set of standard- and low-dose CT scans, resulting in a final model that separately performed well on both types of scans. Unlike previous studies that extract and select a set of slices, our method offers a selection-independent approach to COPD detection and looks at the entire 3D CT volume. We externally validated our model in two large multicenter cohorts, using a variety of scanners. Despite marked differences in the image acquisition protocols of the three cohorts evaluated, COPDxNet showed reliable generalization in both standard- and low-dose chest CT scans.

Another major strength of COPDxNet lies in its lightweight CNN architecture, which has just 5.4 million parameters, significantly lower than many current state-of-the-art architectures. To put this into context, even a relatively simple ResNet-50 architecture has 26 million trainable parameters, whereas more complex variants such as ResNet-152 can have over 60 million parameters.^36^ Similarly, Inception V4 has 48 million parameters, VGG-16 has 138.4 million parameters, and an average vision transformer (ViT) has 86 million parameters.^37^ This lightweight architecture offers a significant advantage for implementation in clinical settings, where computational resources are often limited. Unlike heavier models that demand GPUs or specialized hardware, COPDxNet offers the potential to run efficiently on standard CPUs without sacrificing performance. As a result, it can generate a patient-level prediction of airflow obstruction in under one minute, supporting rapid decision-making in clinical settings.

Our study also has some limitations. Individuals with significant interstitial lung disease were excluded from COPDGene and SPIROMICS; therefore, the performance of COPDxNet remains to be tested in individuals with co-existing lung fibrosis. However, substantial numbers of individuals in each cohort did have interstitial lung abnormalities.^38,39^ Both our derivation and validation cohorts comprised current and former smokers, who are at high risk for COPD; thus, the model should be evaluated in general population cohorts before being applied to identify COPD in non-smokers. Our attempt to mitigate this concern by validating our model in the NLST cohort found relatively lower performance. This difference may be partly explained by the fact that NLST used pre-BD spirometry to assess airflow obstruction, and the scans had a significantly higher slice thickness. Also unlike COPDGene and SPIROMICS, the NLST participants were not coached to TLC before acquiring the inspiratory chest CT.

In conclusion, we demonstrated the effectiveness of a single lightweight 3D CNN, COPDxNet, to detect COPD on both standard and low-dose chest CT scans. COPDxNet has a significantly lower number of parameters compared to other CNN architectures for this indication, making it better-suited for deployment in clinical care, where computational resources are limited.

## FUNDING

This work was supported by NHLBI R01 HL151421 (SPB & AN), NHLBI K01 HL163249 (SB), NHLBI U01 HL089897 and U01 HL089856, and NIH contract 75N92023D00011. COPDGene is also supported by the COPD Foundation through contributions made to an Industry Advisory Board that has included AstraZeneca, Bayer Pharmaceuticals, Boehringer Ingelheim, Genentech, GlaxoSmithKline, Novartis, Pfizer, and Sunovion.

The authors also thank the SPIROMICS participants and participating physicians, investigators, study coordinators, and staff for making this research possible. More information about the study and how to access SPIROMICS data is available at www.spiromics.org. The authors would like to acknowledge the Collaborative Studies Coordinating Center (https://sites.cscc.unc.edu/cscc/), BioSpecimen Processing Facility (http://bsp.web.unc.edu/), and the Alexis Lab and Biospecimen Repository at The University of North Carolina at Chapel Hill for data coordination, sample processing, storage, and sample disbursements.

We would like to acknowledge the following current and former investigators: Neil E Alexis, MD; Wayne H Anderson, PhD; Mehrdad Arjomandi, MD; Igor Barjaktarevic, MD, PhD; R Graham Barr, MD, DrPH; Patricia Basta, PhD; Lori A Bateman, MS; Christina Bellinger, MD; Surya P Bhatt, MD; Eugene R Bleecker, MD; Jessica Bon, MD; Richard C Boucher, MD; Russell P Bowler, MD, PhD; Russell G Buhr, MD, PhD; Stephanie A Christenson, MD; Alejandro P Comellas, MD; Christopher B Cooper, MD, PhD; David J Couper, PhD; Gerard J Criner, MD; Ronald G Crystal, MD; Jeffrey L Curtis, MD; Claire M Doerschuk, MD; Mark T Dransfield, MD; M Bradley Drummond, MD; Christine M Freeman, PhD; Craig Galban, PhD; Katherine Gershner, DO; MeiLan K Han, MD, MS; Nadia N Hansel, MD, MPH; Annette T Hastie, PhD; Eric A Hoffman, PhD; Yvonne J Huang, MD; Robert J Kaner, MD; Richard E Kanner, MD; Joel Kaufman, MD; Mehmet Kesimer, PhD; Eric C Kleerup, MD; Jerry A Krishnan, MD, PhD; Wassim W Labaki, MD; Lisa M LaVange, PhD; Stephen C Lazarus, MD; Nathaniel Marchetti, DO; Fernando J Martinez, MD, MS; Merry-Lynn McDonald, PhD; Deborah A Meyers, PhD; Wendy C Moore, MD; John D Newell Jr, MD; Elizabeth C Oelsner, MD, MPH; Jill Ohar, MD; Wanda K O’Neal, PhD; Claudia Onofrei, MD; Victor E Ortega, MD, PhD; Robert Paine, III, MD; Laura Paulin, MD, MHS; Stephen P Peters, MD, PhD; Cheryl Pirozzi, MD; Nirupama Putcha, MD, MHS; Sanjeev Raman, MBBS, MD; Joseph M Reinhardt, PhD; Stephen I Rennard, MD; Donald P Tashkin, MD; J Michael Wells, MD; Robert A Wise, MD; and Prescott G Woodruff, MD, MPH. The project officers from the Lung Division of the National Heart, Lung, and Blood Institute were Lisa Postow, PhD, Tom Hu, PhD, Lisa Viviano, BSN, and Christine Cochrane; SPIROMICS was supported by contracts from the NIH/NHLBI (HHSN268200900013C, HHSN268200900014C, HHSN268200900015C, HHSN268200900016C, HHSN268200900017C, HHSN268200900018C, HHSN268200900019C, HHSN268200900020C, 75N92024D00012), grants from the NIH/NHLBI (U01HL137880, U24HL141762, R01HL182622, R01HL144718, and R01HL093081), and supplemented by contributions made through the Foundation for the NIH and the COPD Foundation from Amgen; AstraZeneca/MedImmune; Bayer; Bellerophon Therapeutics; Boehringer-Ingelheim Pharmaceuticals, Inc.; Bristol Myers Squibb; Chiesi Farmaceutici S.p.A.; Forest Research Institute, Inc.; Genentech; GlaxoSmithKline; Grifols Therapeutics, Inc.; Ikaria, Inc.; MGC Diagnostics; Novartis Pharmaceuticals Corporation;

Nycomed GmbH; Polarean; ProterixBio; Regeneron Pharmaceuticals, Inc.; Sanofi; Sunovion; Takeda Pharmaceutical Company; Theravance Biopharma; Verona; and Mylan/Viatris.

NLST was supported by the National Institutes of Health (grants U01 CA079778 and U01 CA080098) and by the Department of Health and Human Services via contracts from the Division of Cancer Prevention, National Cancer Institute, and grants to the American College of Radiology Imaging Network and Brown University under a cooperative agreement with the Cancer Imaging Program, Division of Cancer Treatment and Diagnosis, National Cancer Institute. Clinical trial registration number NCT 0047385.

## Data Availability

All data produced are available online at COPDGene, SPIROMICS, and NLST websites.

## REFERENCES

1 Halpin, D. M. G. et al. Global Initiative for the Diagnosis, Management, and Prevention of Chronic Obstructive Lung Disease. The 2020 GOLD Science Committee Report on COVID- 19 and Chronic Obstructive Pulmonary Disease. Am J Respir Crit Care Med 203, 24–36 (2021).

2 Adeloye, D. et al. Global, regional, and national prevalence of, and risk factors for, chronic obstructive pulmonary disease (COPD) in 2019: a systematic review and modelling analysis. The Lancet Respiratory Medicine 10, 447–458 (2022).

3 Metrics, I. Evaluation, Global Burden of Disease Collaborative Network. Global Burden of Disease Study 2016 (GBD 2016) Results. 2017. *Institute for Health Metrics and Evaluation Seattle* (2019).

4 Soriano, J. B. et al. Prevalence and attributable health burden of chronic respiratory diseases, 1990–2017: a systematic analysis for the Global Burden of Disease Study 2017. The Lancet Respiratory Medicine **8**, 585–596 (2020).

5 Sullivan, J. et al. National and State Estimates of COPD Morbidity and Mortality — United States, 2014-2015. J COPD F 5, 324–333 (2018).

6 Bhatt, S. P. et al. Chronic obstructive pulmonary disease: hiding in plain sight, a Statement from the COPD Foundation Medical and Scientific Advisory Committee. The Lancet Respiratory Medicine 11, 1041–1043 (2023).

7 Zafari, Z., Li, S., Eakin, M. N., Bellanger, M. & Reed, R. M. Projecting Long-term Health and Economic Burden of COPD in the United States. Chest 159, 1400–1410 (2021).

8 Siddharthan, T. et al. Discriminative Accuracy of Chronic Obstructive Pulmonary Disease Screening Instruments in 3 Low- and Middle-Income Country Settings. JAMA 327, 151 (2022).

9 Martinez, C. H. et al. Undiagnosed Obstructive Lung Disease in the United States. Associated Factors and Long-term Mortality. Annals ATS 12, 1788–1795 (2015).

10 Diab, N. et al. Underdiagnosis and Overdiagnosis of Chronic Obstructive Pulmonary Disease. Am J Respir Crit Care Med 198, 1130–1139 (2018).

11 Baldomero, A. K. et al. Beyond Access: Factors Associated With Spirometry Underutilization Among Patients With a Diagnosis of COPD in Urban Tertiary Care Centers. Chronic Obstr Pulm Dis 9, 538–548 (2022).

12 Jain, V. V. et al. Misdiagnosis Among Frequent Exacerbators of Clinically Diagnosed Asthma and COPD in Absence of Confirmation of Airflow Obstruction. Lung 193, 505–512 (2015).

13 Smith-Bindman, R. et al. Trends in Use of Medical Imaging in US Health Care Systems and in Ontario, Canada, 2000-2016. JAMA **322**, 843 (2019).

14 Sarma, A. et al. Radiation and Chest CT Scan Examinations. Chest 142, 750–760 (2012).

15 Berrington De González, A. Projected Cancer Risks From Computed Tomographic Scans Performed in the United States in 2007. Arch Intern Med 169, 2071 (2009).

16 González, G. et al. Disease Staging and Prognosis in Smokers Using Deep Learning in Chest Computed Tomography. Am J Respir Crit Care Med 197, 193–203 (2018).

17 Tang, L. Y. W. et al. Towards large-scale case-finding: training and validation of residual networks for detection of chronic obstructive pulmonary disease using low-dose CT. The Lancet Digital Health 2, e259–e267 (2020).

18 Vestbo, J. et al. Evaluation of COPD Longitudinally to Identify Predictive Surrogate End- points (ECLIPSE). European Respiratory Journal (2008) doi:10.1183/09031936.00111707.

19 Amudala Puchakayala, P. R., et al. Radiomics for Improved Detection of Chronic Obstructive Pulmonary Disease in Low-Dose and Standard-Dose Chest CT Scans. Radiology 307, e222998 (2023).

20 Regan, E. A. et al. Genetic epidemiology of COPD (COPDGene) study design. COPD: Journal of Chronic Obstructive Pulmonary Disease (2010) doi:10.3109/15412550903499522.

21 Collins, G. S., Reitsma, J. B., Altman, D. G. & Moons, K. G. M. Transparent reporting of a multivariable prediction model for individual prognosis or diagnosis (TRIPOD) the TRIPOD statement. Circulation (2015) doi:10.1161/CIRCULATIONAHA.114.014508.

22 Collins, G. S. et al. TRIPOD+AI statement: updated guidance for reporting clinical prediction models that use regression or machine learning methods. BMJ e078378 (2024) doi:10.1136/bmj-2023-078378.

23 Bhatt, S. P. et al. Imaging Advances in Chronic Obstructive Pulmonary Disease. Insights from the Genetic Epidemiology of Chronic Obstructive Pulmonary Disease (COPDGene) Study. Am J Respir Crit Care Med 199, 286–301 (2019).

24 Couper, D. et al. Design of the subpopulations and intermediate outcomes in copd study (SPIROMICS). Thorax (2014) doi:10.1136/thoraxjnl-2013-203897.

25 Sieren, J. P. et al. SPIROMICS Protocol for Multicenter Quantitative Computed Tomography to Phenotype the Lungs. Am J Respir Crit Care Med 194, 794–806 (2016).

26. 26 National Lung Screening Trial Research Team. The National Lung Screening Trial: Overview and Study Design. *Radiology* 258, 243–253 (2011).

27 Fluss, R., Faraggi, D. & Reiser, B. Estimation of the Youden Index and its Associated Cutoff Point. Biometrical J 47, 458–472 (2005).

28 Guo, C., Pleiss, G., Sun, Y. & Weinberger, K. Q. On calibration of modern neural networks. in Proceedings of the Machine Learning Research vol. 70 1321–1330 (2017).

29 Schroeder, J. D. et al. Prediction of Obstructive Lung Disease from Chest Radiographs via Deep Learning Trained on Pulmonary Function Data. Int J Chron Obstruct Pulmon Dis 15, 3455–3466 (2020).

30 Du, R. et al. Identification of COPD From Multi-View Snapshots of 3D Lung Airway Tree via Deep CNN. IEEE Access 8, 38907–38919 (2020).

31 Ho, T. T. et al. A 3D-CNN model with CT-based parametric response mapping for classifying COPD subjects. Sci Rep 11, 34 (2021).

32 Jonas, D. E. et al. Screening for Lung Cancer With Low-Dose Computed Tomography: Updated Evidence Report and Systematic Review for the US Preventive Services Task Force. JAMA 325, 971 (2021).

33 Meza, R. et al. Evaluation of the Benefits and Harms of Lung Cancer Screening With Low-Dose Computed Tomography: Modeling Study for the US Preventive Services Task Force. JAMA 325, 988 (2021).

34 Hammond, E. et al. Comparison of low- and ultralow-dose computed tomography protocols for quantitative lung and airway assessment. Med. Phys. 44, 4747–4757 (2017).

35 Hatt, C. R. et al. Comparison of CT Lung Density Measurements between Standard Full-Dose and Reduced-Dose Protocols. Radiology: Cardiothoracic Imaging 3, e200503 (2021).

36 Tan, M. & Le, Q. Efficientnet: Rethinking model scaling for convolutional neural networks. in 6105–6114 (PMLR, 2019).

37 Khan, S. et al. Transformers in Vision: A Survey. ACM Comput. Surv. 54, 1–41 (2022).

38 Washko, G. R. et al. Lung Volumes and Emphysema in Smokers with Interstitial Lung Abnormalities. N Engl J Med 364, 897–906 (2011).

39 Baddour, N. A. et al. Air Pollution Exposure and Interstitial Lung Features in SPIROMICS Participants with Chronic Obstructive Pulmonary Disease. Ann Am Thorac Soc 21, 1251– 1260 (2024).

