## Supplementary Appendix for "Light Convolutional Neural Network to Detect Chronic Obstructive Pulmonary Disease (COPDxNet): A Multicenter Model Development and External Validation Study"

**Supplementary Appendix Light Convolutional Neural Network for Detecting Chronic Obstructive Pulmonary Disease (COPDxNet): A Multicenter Model Development and External Validation**

AKM Shahariar Azad Rabby, M.S.,^1,2,*^ Muhammad F. A. Chaudhary, Ph.D.,^1,3,*^ Pratim Saha, B.S.,^1,2^ Venkata Sthanam, M.S.,^1,4^ Arie Nakhmani, Ph.D.,^1,4^ Chengcui Zhang, Ph.D.,^2^ R. Graham Barr, M.D.,^5^ Jessica Bon, M.D.,^6^ Christopher B. Cooper, M.D.,^7^ Jeffrey L. Curtis, M.D.,^8^ Eric A. Hoffman, Ph.D.,^9^ Robert Paine, III, M.D.,^10^ Abhilash Kizhakke Puliyakote, Ph.D.,^9^ Joyce D. Schroeder, M.D.,^11^ Jessica C. Sieren, Ph.D.,^9^ Benjamin M. Smith, M.D., M.S.,^5^ Prescott Woodruff, M.D.,^12^ Joseph M. Reinhardt, Ph.D.,^13^ Surya P. Bhatt, M.D., M.S.P.H,^1,3,†^ and Sandeep Bodduluri, Ph.D.,^1,3,†^

1. Center for Lung Analytics and Imaging Research (CLAIR), The University of Alabama at Birmingham, Birmingham, AL, 35294.
2. Department of Computer Science, The University of Alabama at Birmingham, AL, 35294;
3. Division of Pulmonary, Allergy and Critical Care Medicine, The University of Alabama at Birmingham, Birmingham, AL, 35294.
4. Department of Electrical and Computer Engineering, The University of Alabama at Birmingham, Birmingham, AL, 35294
5. Department of Epidemiology, Mailman School of Public Health, Columbia University, New York, NY, 10032.
6. Division of Pulmonary, Critical Care, Allergy and Immunology, School of Medicine, Wake Forest University, Winston-Salem, NC, 27109.
7. Department of Physiology, David Geffen School of Medicine at UCLA, Los Angeles, CA, 90095.
8. Division of Pulmonary and Critical Care Medicine, University of Michigan Health System, Ann Arbor, MI, 48109.
9. Department of Radiology, University of Iowa Roy J. and Lucille A. Carver College of Medicine, Iowa City, IA, 52242.
10. Division of Respiratory, Critical Care and Occupational Pulmonary Medicine, University of Utah, Salt Lake City, UT, 84112.
11. Department of Radiology, Mayo Clinic, Rochester, MN, 55905.
12. Division of Pulmonary, Critical Care, Allergy and Sleep Medicine, Department of Medicine, University of California, San Francisco, CA, 94115.
13. The Roy J. Carver Department of Biomedical Engineering, The University of Iowa, Iowa City, IA, 52246.

*Equal Contribution

^†^Co-senior Authors

**Corresponding Author:** Sandeep Bodduluri, Ph.D., Division of Pulmonary, Allergy and Critical Care Medicine, The University of Alabama at Birmingham, 930 20^th^ St S (Room 240), Birmingham, AL 35233.. Phone: 205-934-5555. Fax: 205-934-6229.

**Funding Sources:** This work was supported by NHLBI R01 HL151421 (SPB & AN), NHLBI K01Hl163249 (SB), NHLBI U01 HL089897 and U01 HL089856 and NIH contract 75N92023D00011. COPDGene is also supported by the COPD Foundation through contributions made to an Industry Advisory Board that has included AstraZeneca, Bayer Pharmaceuticals, Boehringer Ingelheim, Genentech, GlaxoSmithKline, Novartis, Pfizer, and Sunovion.

**Manuscript Word Count**: 2704 / 3500

**Supplementary Methods**

**COPDGene Inclusion and Exclusion Criteria**

The COPDGene study enrolled adults aged 45 to 80 years with a smoking history of at least 10 pack-years. The study enrolled 10,305 individuals of non-Hispanic white or African American ethnicity, who were capable of providing informed consent and undergoing imaging and pulmonary function tests. Eligibility criteria are listed in **Table S1**.^1^ Spirometry and chest computed tomography (CT) scans were acquired at 21 clinical centers across the United States. Several CT scanner types were used to acquire chest CT scans including scanners from Siemens, General Electric (GE), and Koninklijke Philips N.V. commonly known as Philips.^1^

**Table S1**: COPDGene inclusion and exclusion criteria.

| **Criteria** | **COPD Cases** | **Smokers without COPD** |
| --- | --- | --- |
| **Inclusion** | | |
| Age | 45–80 years | 45–80 years |
| Smoking History | ≥ 10 pack-years | ≥ 10 pack-years |
| Lung Function | GOLD stages 2, 3, or 4 (post-bronchodilator FEV_1_/FVC < 0.70 and FEV_1_ < 80% predicted) | Post-bronchodilator FEV_1_/FVC ≥ 0.70 and FEV_1_ > 80% predicted |
| Ethnicity | Non-Hispanic White or African American | Non-Hispanic White or African American |
| **Exclusion** | | |
| Respiratory Diseases | Concomitant respiratory disorder other than COPD or asthma (e.g., bronchiectasis, interstitial lung disease) | Physician-diagnosed respiratory disease other than COPD or asthma |
| Surgical History | Lung surgery (lobe removal, lung volume reduction, or transplantation) | Lung surgery (lobe removal, lung volume reduction, or transplantation) |
| Cancer | Lung cancer or uncontrolled cancer (e.g., ongoing chemo/radiation, narcotics for pain, metastasis) | Uncontrolled cancer (e.g., ongoing chemo/radiation, narcotics for pain, metastasis) |
| Radiation | Chest radiation therapy (excluding breast cancer) | Chest radiation therapy |
| Recent Medications | Antibiotics or systemic steroids for COPD exacerbation or respiratory infection within the past month | Antibiotics or systemic steroids for respiratory infection within the past month |
| Device Interference | Metal objects (pacemakers, defibrillators, prosthetic heart valves, shoulder prostheses) | Metal objects (pacemakers, defibrillators, prosthetic heart valves, shoulder prostheses) |
| Specific Conditions | Recent surgery, heart attack, or detached retina (within 3 months) | Recent surgery, heart attack, or detached retina (within 3 months) |
| Study Participation | Participation in specific studies (e.g., ECLIPSE, COPDGene, or others) | Participation in specific studies (e.g., ECLIPSE, COPDGene, or others) |
| Family Enrollment | First/second-degree relative in COPDGene | First/second-degree relative in COPDGene |
| Racial Category | Subjects indicating more than one racial category | Subjects indicating more than one racial category |

**Model Architecture Details**

COPDxNet is a low-parameter, efficient, 3D convolutional neural network (CNN) developed for detecting chronic obstructive pulmonary disease (COPD) from inspiratory chest CT scans. We developed COPDxNet using the TensorFlow framework for binary classification. COPDxNet processes 3D CT volumes through a sequence of convolutional blocks, fully-connected layers, and an output layer, ultimately generating a probability score that indicates the likelihood of COPD. The input to the network is a tensor of size (N, 128, 128, 64, 1), where *N* is the batch size defined during training, and each volume has a single channel.

The model architecture consists of a series of convolutional blocks, each with a 3D convolutional layer, rectified linear unit (ReLU) activation, batch normalization, max pooling, and dropout to prevent overfitting. The blocks progressively increase the number of filters (64, 128, 256) to capture multiresolution features at different scales. After the convolutional layers, the model flattens the feature maps and passes them through a dense layer with 512 units, applying ReLU activation and further dropout for regularization. Finally, the output layer with a single sigmoid-activated unit provides a probability score, classifying the presence or absence of COPD.

**Data Augmentation Techniques**

To improve model generalization, we used several data augmentation techniques that were applied exclusively to the training data. The augmentations included random rotations within a range of -5° to 5°. The model was compiled using binary cross-entropy loss and the Adam optimizer, with an initial learning rate of 0.0001, decaying exponentially by 0.96 every 100,000 steps.

**Supplementary Results**

| **Table S2**: Participant characteristics for the low-dose chest CT dataset from the NLST cohort.^2^ | |
| --- | --- |
|  | **NLST (Low-Dose)** |
|  | *n* = 7893 |
| **Age, years** | 61.7 (5.1) |
| **Sex** |  |
| **Male, n (%)** | 4376 (55.4) |
| **Female, n (%)** | 3517 (44.6) |
| **Race** |  |
| **White, n (%)** | 7295 (92.4) |
| **Black, n (%)** | 448 (5.7) |
| **Smoking history** | 28.8 (6.2) |
| **Current smokers, n (%)** | 3957 (50.1) |

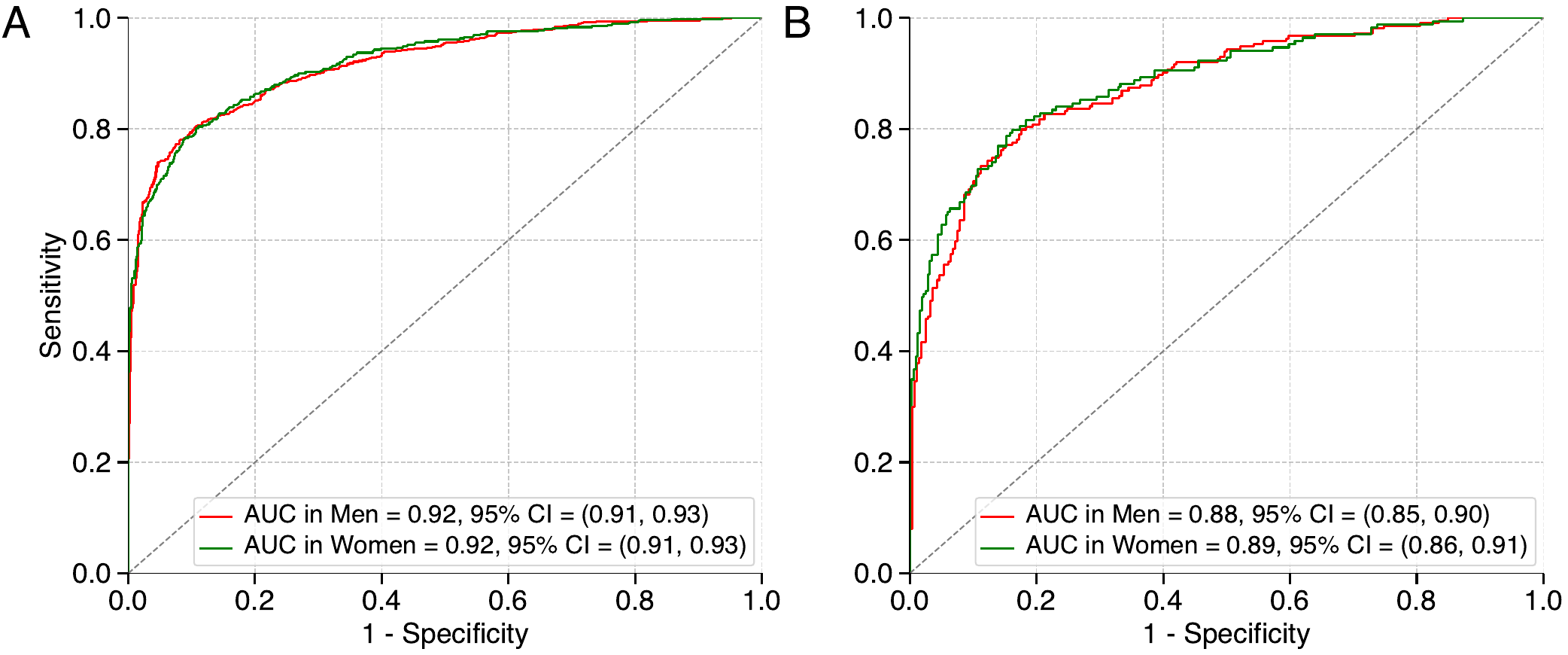

**Figure S1**: ROC curve analysis for assessing COPDxNet performance, stratified by sex, on standard-dose (Panel A) and low-dose (Panel B) chest CT scans.

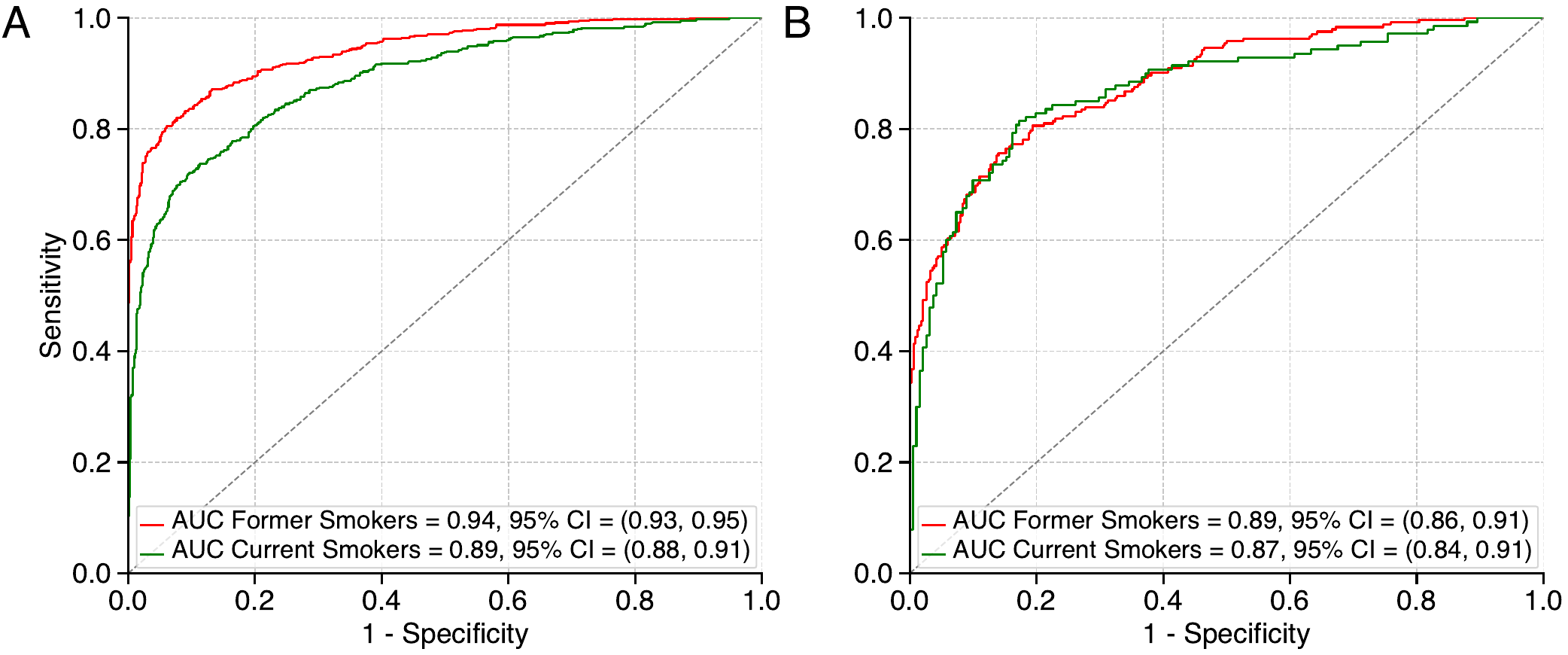

**Figure S2**: Performance evaluation of COPDxNet in current and former smokers with standard (Panel A) and low-dose (Panel B) CT scans.

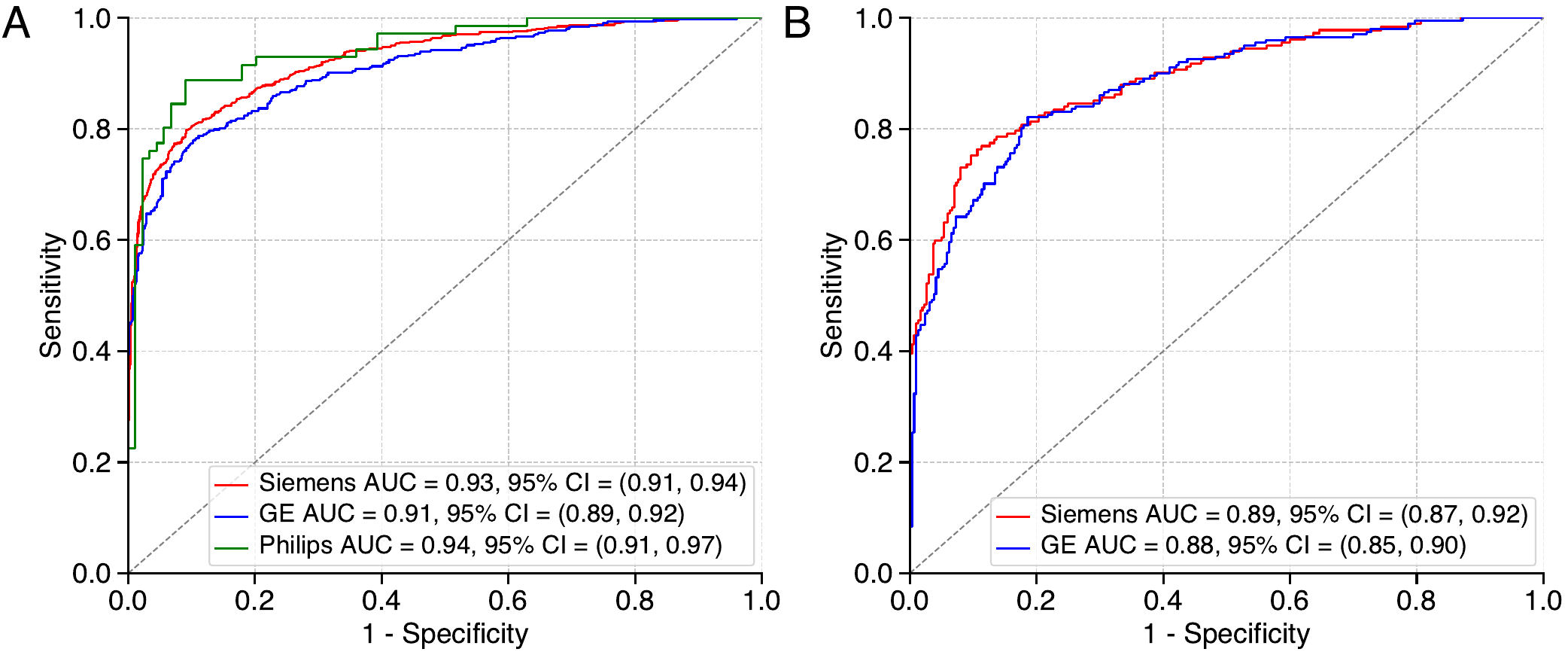

**Figure S3**: COPDxNet performance by scanner type (Siemens, GE, and Philips) on standard (Panel A) and low-dose (Panel B) scans. For low-dose CT scans, Philips scanner type had no individuals with COPD at visit 3, and hence was excluded from analysis shown in Panel B.

**References**

1.	Regan, E. A. *et al.* Genetic epidemiology of COPD (COPDGene) study design. *COPD: Journal of Chronic Obstructive Pulmonary Disease* (2010) doi:10.3109/15412550903499522.

2.	National Lung Screening Trial Research Team. The National Lung Screening Trial: Overview and Study Design. *Radiology* **258**, 243–253 (2011).
